# Impairment of dual-task gait dynamics in older adults with mild cognitive impairment: Relationships to neuropsychological status, fitness and brain morphology

**DOI:** 10.1101/19005249

**Authors:** Tess C Hawkins, Rebecca Samuel, Maria A Fiatarone Singh, Nicola Gates, Guy C Wilson, Nidhi Jain, Jacinda Meiklejohn, Henry Brodaty, Wei Wen, Nalin Singh, Bernhard T Baune, Chao Suo, Michael K Baker, Nasim Foroughi, Yi Wang, Perminder S Sachdev, Michael J Valenzuela, Jeffrey M Hausdorff, Yorgi Mavros

## Abstract

**Background:** Individuals with Mild Cognitive Impairment (MCI) have more gait variability under dual-task conditions than cognitively healthy adults. However, characteristics associated with this susceptibility of gait to dual-task stress are unknown.

**Methods:** Testing was performed at baseline in the Study of Mental And Resistance Training (SMART). Ninety-three adults with MCI (age 70±6.8 years; 66.6% female) performed a single- and dual-task walk (cognitive distractor=letter fluency), in random order. Linear and non-linear gait variability were measured using force-sensitive insoles. Cognitive performance during dual-tasking was assessed by the number of correct words vocalized. Cognitive function, brain Magnetic Resonance Imaging (MRI), muscle strength, aerobic capacity, body composition, physical and psychosocial function were also assessed as potential correlates of gait dynamics.

**Results:** Gait dynamics worsened during dual-tasking, with decrements in both stride time variability (p<0.001) and detrended fluctuation analysis (DFA) (p=0.001). Lower aerobic capacity and thinner posterior cingulate cortex were associated with greater decrements in DFA (p<0.05). Smaller hippocampal volume, worse psychological well-being and poorer static balance were associated with greater decrements in stride time variability (p<0.05). By contrast, cognitive performance did not change under dual-task conditions compared to seated testing (p=0.13).

**Conclusions:** Under dual-task conditions, participants with MCI preserved their cognitive performance at the expense of gait stability. Decrements in dual-tasking gait were associated with lower aerobic fitness, balance, psychological well-being, and brain volume in cognitively-relevant areas of the posterior cingulate and hippocampus, all potentially modifiable characteristics. Trials of targeted interventions are needed to determine the potential plasticity of gait variability in high-risk cohorts.

## INTRODUCTION

The prevalence of gait abnormalities increases with age, affecting up to 35% of community-dwelling older adults1. Gait abnormalities are characterized by a more variable gait pattern, leaving affected individuals with an increased risk of falls^2^ and reduced mobility^3^. Adults with mild cognitive impairment (MCI), an intermediate stage between normal cognition and dementia, are more likely to have greater gait variability^4^ and consequently, double the risk of injurious and multiple falls compared to cognitively normal adults^5^.

Dual-tasking is a sensitive method used to investigate interactions between gait variability and cognitive function^6^, with worse gait performance under dual-task conditions associated with an increased fall risk amongst older adults^78^. Dual-tasking impairs gait in individuals with deficits in cognitive function, including MCI and Alzheimer’s disease^9^. However, studies evaluating dual-task associations between cognition and gait variability in older adults with MCI are limited. Furthermore, imaging studies of brain regions and their associations with gait variability have mainly focussed on single-task gait variability, or the effects of dual-tasking on gait speed^10-12^ rather than more comprehensive assessments of gait variability or cognitive performance under dual-task stress. Finally, identification of modifiable characteristics associated with gait dynamics under dual-task conditions could lead to targeted interventions to improve gait dynamics and ultimately reduce falls in older adults with MCI and other vulnerable cohorts. Therefore, the aim of the present study was to determine the effects of dual-tasking on gait dynamics and cognitive performance in older adults with MCI, as well as to identify relevant physical, psychosocial, and structural brain characteristics associated with decrements during dual-tasking (dual-task cost). We hypothesized that there would be a worsening of *both* gait and the performance of the secondary cognitive task during the dual-task condition, and that these reductions would be associated with lower strength, aerobic capacity, functional performance, psychosocial function and smaller hippocampal volume and posterior cingulate cortex thickness. These factors were selected *a priori* due to their known decrements in individuals with cognitive impairment, frailty, or elevated falls risk^13^.

## METHODS

The complete study protocol for the Study of Mental and Resistance Training (SMART) has been published^14^, with primary^15^ and secondary outcomes published^16,17^. The study was approved by the Royal Prince Alfred Human Research Ethics Committee (X08-0064) on 24th of April, 2008, and written informed consent was obtained from all participants. The study was prospectively registered with the Australia New Zealand Clinical Trials Registry (ACTRN12608000489392). A cross-sectional analysis of baseline data from the SMART trial was used for this investigation.

### Participants

One hundred community-dwelling older adults with MCI (Peterson criteria^18^) were recruited. Two participants could not wear gait monitors due to fused toes, while technical issues including incomplete data recording, precluded full gait data in five others. Thus, gait dynamics data were available for 93 participants. All participants were assessed at The University of Sydney, New South Wales, Australia.

### Assessment of cognitive function

The primary outcome of the SMART trial was global cognition assessed using the Alzheimer’s Disease Assessment Scale - Cognition (ADAS-Cog). Global, executive, and memory cognitive domains were also calculated for the 93 participants with available gait variability data. Additional details can be found in the Supplementary methods.

### Assessment of letter fluency at rest

Letter fluency, a subcategory of verbal fluency, was assessed using the COWAT^19^ to measure cognitive performance at rest. All participants were tested in a private room with a single research assistant, in a resting, non-fasting state at a similar mid-morning time. Participants were instructed to verbalize as many words as possible beginning with the letter “F” in 1 minute (F_SINGLE_), excluding proper nouns, repeated words, and variations of the same word using a prefix or suffix (e.g., foam and foaming). A score was calculated by totaling the number of admissible words written down by the research assistant.

### Assessment of gait dynamics

Gait dynamics were assessed similarly to methods previously described^20,21^. Briefly, force-sensitive insoles were placed in the participants’ shoes to measure the force applied to the ground during a 2-minute period of ambulation at habitual pace using any assistive devices needed. Participants were instructed to wear low-heeled shoes, comfortable clothing and visual and hearing aids if required.

Gait assessment was performed under two conditions, ‘single-task’ (undistracted walking) and ‘dual-task’ (distracted walking), in a randomized order. Participants were not provided with any instruction regarding task priority (i.e, cognitive or gait performance), and were told to perform as well as possible for both tasks. Stride time variability (coefficient of variation, CV) and detrended fluctuation analyses fractal scaling exponent (DFA^22,23^ were calculated for both single-task [(CV_SINGLE_) and (DFA_SINGLE_)] and dual-task [(CV_DUAL_) and (DFA_DUAL_)] conditions. To assess stride-to-stride variability and arrhythmicity of gait, the coefficient of variation (CV) in each participant’s stride time was calculated using the formula CV = (Standard Deviation / Mean) * 100, with lower CV indicating more stable gait. To quantify how the dynamics of stride times fluctuate and change over time, we applied detrended fluctuation analysis (DFA) to each participant’s sequence of stride times. In general, physiologically healthy systems have fractal scaling indices between 0.8 and 1.0, with lower values (closer to 0.5) indicating a less healthy state^24^.

The secondary (cognitive distractor) task was the COWAT test, with the number of correct “F words” counted in the first minute of walking (F_DUAL_). The letter “F” was selected to remain consistent with the seated COWAT test described above (F_SINGLE_). The words were recorded using a portable recorder (Samsung YP-U3, Samsung Electronics Co., South Korea) and the recording was subsequently reviewed by the research assistant who counted the number of admissible words during the first and second minute. The number of correct words in the first minute only of the dual-task gait condition (F_DUAL_) was compared to the number of correct words during the F_SINGLE_ condition. The order was not randomized for the assessment of “F” words, with the dual-task condition performed 1 week after the seated COWAT. The ‘dual-task cost’ for CV, DFA and F words were calculated (CV_COST_= CV_DUAL_ – CV_SINGLE_; DFA_COST_= DFA_DUAL_ – DFA_SINGLE_; F_COST_ = F_DUAL_ – F_SINGLE_).

### Assessment of Neuroimaging Outcomes

Details of the neuroimaging assessment outcomes have been published^14,17^. Briefly, MRI data were acquired on a 3.0-Tesla Philips Achieva System (Amsterdam, The Netherlands). Brain structure was assessed using a T1-weighted whole-brain scan with isotropic voxel 1 mm^3^ (sequence: T1TFE; TR/TE: 6.39/2.9 ms; slice thickness 1.0 mm without gap; in-plane resolution 1 × 1 mm). Left, right hippocampal volume was manually traced, using ANALYZE (version 10, Mayo Clinic) following the published protocol^25^ For cortical thickness measurement, FreeSurfer (v5.1.0, http://freesurfer.net) was applied to calculate the cortical thickness at whole brain level. Then, the posterior cingulate mean cortical thickness was generated using the standard FreeSurfer parcellation (Desikan-Killiany Atlas)^26^.

### Assessment of peak strength

Maximal strength testing was assessed via the one-repetition maximum (1RM) on Keiser pneumatic resistance machines (Keiser Sports Health Equipment, Ltd., Fresno, CA, USA). Participants’ 1RMs were determined on the leg press, knee extension, hip abduction, chest press and seated row machines. One RM tests were performed twice, one week apart, with the best performance used.

### Assessment of peak aerobic capacity

Aerobic capacity (VO_2_peak) was determined via indirect calorimetry during a physician-administered, graded treadmill walking test to volitional fatigue. Methods and data handling have been previously published^16^.

### Measures of physical function

Details of the physical function assessments have been published^14^. Static balance was assessed on one attempt using six different positions (wide stance, narrow stance, semi-tandem stance, tandem stance, one leg with eyes open and one leg with eyes closed). Dynamic balance was assessed using the time taken to forward tandem walk over a 3-meter marked course. Habitual and maximal gait velocities were assessed over two metres using an Ultra-timer (Raymar, Oxfordshire, UK). Lower extremity function and power was assessed using time to complete 5 sit-to-stands and stair climb power. Stair climb power was calculated using the following formula P (watts) = (M × D) × 9.8/t Where: M = Body mass (kg), D= Vertical distance (m), D = vertical height of the staircase, and t = Time (s). The 6-minute walk test (6MWT) was assessed twice, at least one week apart, with the furthest distance walked used. Further detail regarding these assessments can be found in the Supplementary methods.

### Anthropometry and Body Composition

Height, naked body mass and waist circumference were measured as the mean of triplicate measures after a 12-hour overnight fast. Waist circumference was measured with Lufkin steel tape measure (W606 PM, Apex Tool Australia Pty Ltd, Albury, NSW, AU), using the International Diabetes Federation (IDF) protocol. Bioelectrical Impedance Analysis (BIA; RJL Systems, Inc., Clinton, MI, USA) was used to evaluate body composition. Whole body skeletal muscle mass (kg) and fat free mass (kg) were calculated using the average resistance and reactance values of three sequential BIA measures. Further details of the assessments are published^14^.

### Measures of Psychosocial Function

Psychosocial function and wellbeing were measured via the Life Satisfaction Scale (LSS), Scale of Psychological Well Being (SPWB), Quality of Life Scale (QOLS), Physical and Mental Health Short-36 (SF-36), Depression Anxiety Stress Scale (DASS 21), Memory Awareness Rating Scale – Memory Functioning Scale (MARS-MF), Duke Social Support Index Scale (DSSIS) and Life Experience Questionnaire (LEQ). Further detail regarding these assessments has been published^14^

### Statistical Methods

Data were inspected for normality. Normally-distributed data are presented as mean±SD and non-normally distributed data presented as median (interquartile range). All CV (CV_SINGLE_, CV_DUAL_ and CV_COST_) variables were log-transformed prior to use in parametric statistics. Sequential linear regression models (adjusted for age and sex) were constructed including potential confounders associated with the number of F words during the COWAT test (F_SINGLE_) and single- and dual-task gait dynamics (CV_SINGLE_ and DFA_SINGLE_). Next, linear mixed models with repeated measures were constructed to determine the effect of dual-tasking on both letter fluency and gait dynamics. The single-task condition (F_SINGLE_, CV_SINGLE_ and DFA_SINGLE_) was entered as time point 1, and the ‘dual-task’ condition (F_DUAL_, CV_DUAL_ and DFA_DUAL_) as time point 2. A compound symmetry covariance matrix was used. Models were adjusted for age and sex, with gait dynamics outcomes further adjusted for the order of the walking condition. Next, linear regression models were constructed to determine variables associated with F_COST_, CV_COST_ and DFA_COST_. Models were adjusted for age, sex, and baseline score of the dependent variable and order of the condition for the gait dynamics assessment. All models involving cognitive performance (including F_SINGLE_ and F_COST_) were further adjusted for education, while models involving hippocampal volume and posterior cingulate cortex thickness were adjusted further adjusted for total intracranial volume. Models involving DFA were also adjusted for the number of medications, as justified below. Statistical significance was assumed at α<0.05. All data were analysed using IBM SPSS (version 24; IBM Corp., Armonk, NY, USA).

## RESULTS

Baseline clinical characteristics of the participants have been published^15^. Data for the available 93 participants (66.6% women), were similar to the overall cohort (p>0.05 for all variables). The average age was 70.0±6.8 years, MMSE score 27.5±1.4, and habitual gait speed 1.21±0.24 m/s, with 17% of participants having a gait speed below 1.0 m/s. Participant characteristics data are presented in Table 1.

**TABLE 1:** Participant characteristics.

| **Characteristics** | **Value** |
| --- | --- |
| *Demographics* |  |
| Age (years) | 70.0 ± 6.8 |
| Sex: Female (%) | 66.7 |
| BMI (kg/ m<sup>2</sup>) | 26.87 ± 4.89 |
| Education (years) | 13 ± 3 |
| Total alcoholic drinks / week (n) | 3 (9) |
| Current smoker (%) | 2.2 |
| Ex-smoker (%) | 38.7 |
| *Health status* |  |
| Medications/day (n) | 4 (4) |
| Number of chronic diseases (n) | 2.9 ± 1.7 |
| Osteoarthritis (%) | 71.0 |
| Hypertension (%) | 40.9 |
| Diabetes (%) | 11.8 |
| Gout (%) | 4.3 |
| Depressive episodes in the past 5 years (n) | 0 (0) |
| Habitual gait speed (m/s) | 1.22 ± 0.24 |

Results reported in mean ± SD or Median (IQR); SD=Standard deviation; IQR=Interquartile
Range; m/s= meters per second; kg/m<sup>2</sup>= kilograms per meter squared; BMI= body mass index

### Factors associated with single task gait dynamics

The number of medications prescribed was inversely associated with DFA_SINGLE_ (r=-0.23, p=0.029), but not CV_SINGLE_ (r=-0.03, p=0.75). There were no associations between age, sex, years of education, smoking status, drinking status or the number of chronic diseases and either index of gait dynamics. Consequently, all analyses with DFA_COST_ as a dependent variable were adjusted for the number of medications.

### Dual-Task Cost

Data are presented in Figure 1. As hypothesized, gait dynamics worsened significantly during dual-tasking, with decrements in performance observed for stride time variability [single-task 2.012 (0.767), dual-task 2.555 (2.227); p<0.0001 and DFA (single-task 0.804±0.151, dual-task 0.745±0.160; p=001). However, contrary to our hypothesis, cognitive performance on the COWAT did not significantly change under dual-task conditions (single-task 13±5 words, dual-task 12±4 words, p=0.13; Figure 1). Changes in letter fluency (F_COST_) were not associated with changes in gait dynamics for either CV_COST_ (r=-0.14, p=0.171) or DFA_COST_ (r=0.03, p=0.746).

**FIGURE 1:**
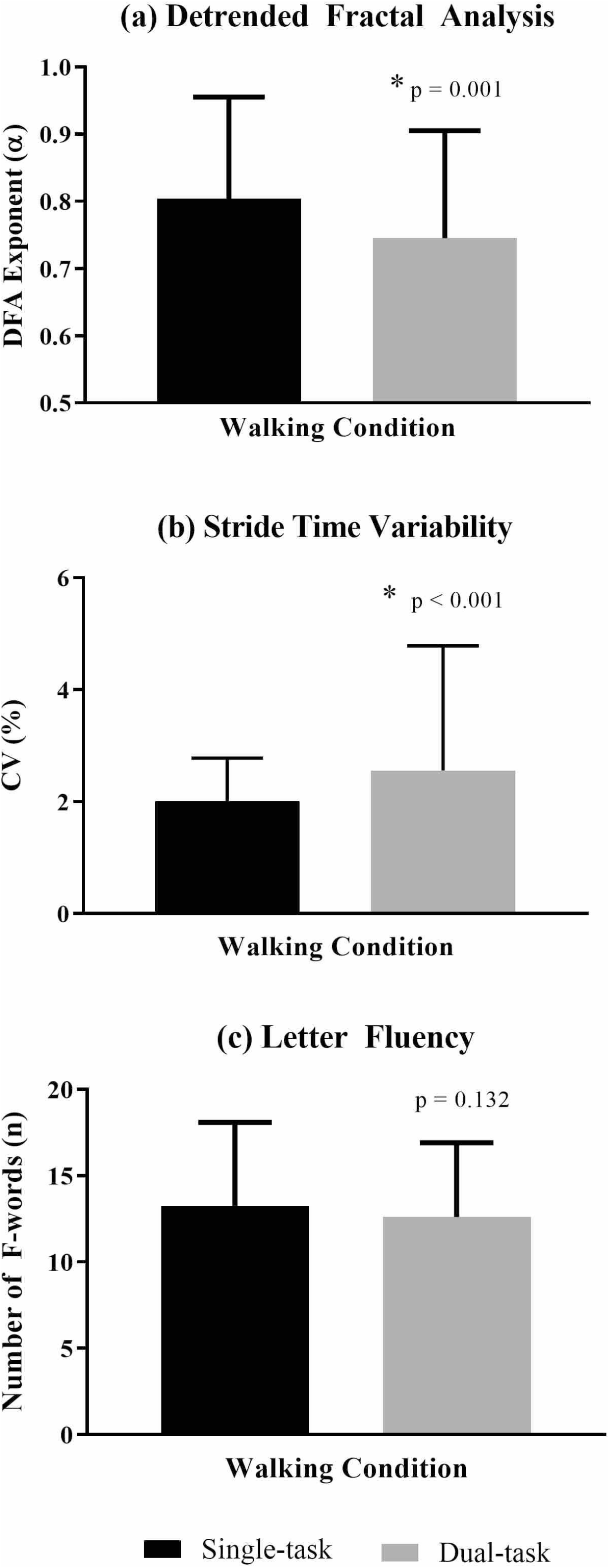
Gait dynamics and cognitive performance under single-task and dual-task walking conditions. Graphed results under single-task and dual-task walking conditions for (a) DFA, (b) Stride time variability (CV) and (c) Letter fluency. Data for (a) and (c) are presented as mean±SD and data for (b) are presented as median (IQR) with all adjusted pairwise. DFA = detrended fluctuation analysis fractal scaling exponent; CV = coefficient of variation. Higher scores are worse for CV. Lower scores are worse for DFA and Letter Fluency.

### Factors associated with changes in stride time variability component of gait during dual-tasking (CV_*COST*_)

For simplicity, significant associations are presented in Table 2. A complete set of results can be seen in the Supplemental material. Contrary to our hypotheses, cognitive performance (executive function, memory, attention and global domains) was not associated with CV_COST_ under dual-task conditions (p>0.05). However, higher (worse) CV_COST_ was associated with smaller left hippocampus volume (r=-0.35, p=0.023) with a similar trend for total hippocampus volume (r=-0.31, p=0.050). Gait dynamics and brain morphology data are presented in Figure 2.

**TABLE 2:**
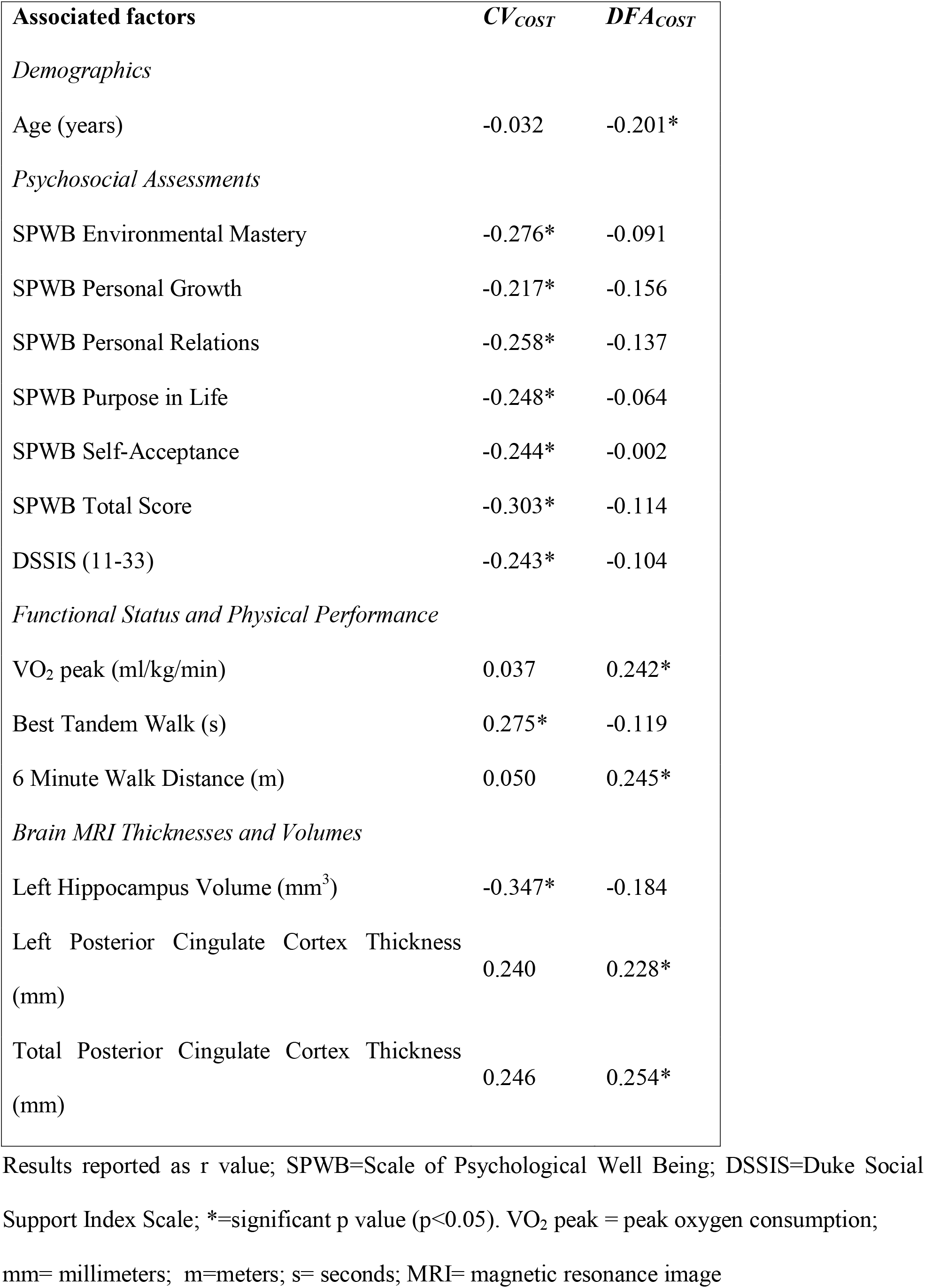
Factors significantly associated with changes in at least one measure of gait variability or dynamics during dual-tasking.

**FIGURE 2:**
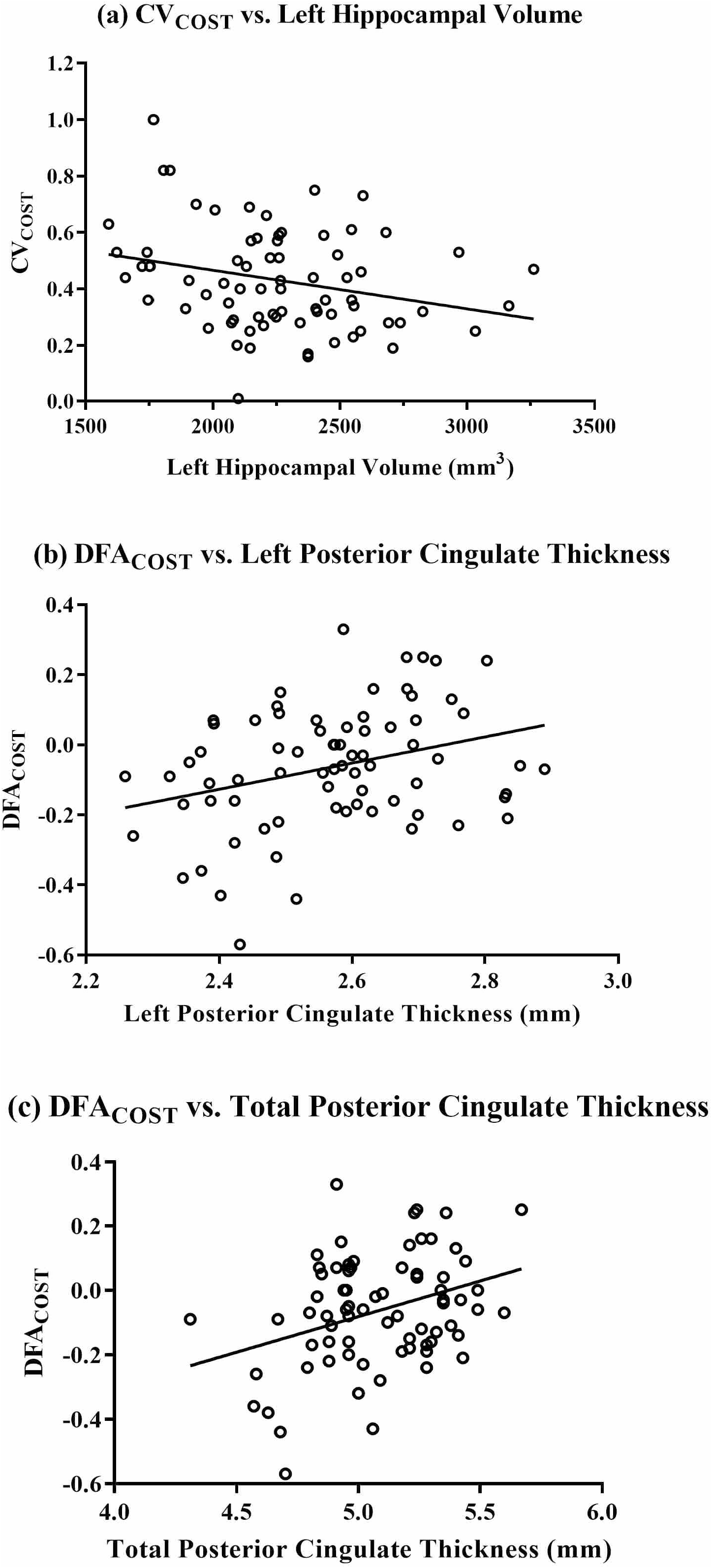
Relationship between gait dynamics and brain morphology Graphed results of gait dynamics and MRI associations for (a) CV_COST_ and left hippocampal volume, (b) DFA_COST_ and left posterior cingulate thickness, and (c) DFA_COST_ and total posterior cingulate thickness. CV_COST_ = the dual-task cost of stride time variability. DFA_COST_ = the dual-task cost of DFA. DFA = detrended fluctuation analysis fractal scaling exponent. CV = coefficient of variation. Cost variables are calculated by subtracting the single-task from the dual-task. Higher cost indicates greater gait decrement during dual-tasking.

Worse dynamic balance (longer tandem walk time) was directly associated with a higher CV_COST_ (r=0.28, p=0.022). However, CV_COST_ was not associated with body composition, aerobic capacity, strength or other measures of functional performance.

As hypothesized, psychological well-being was inversely associated with CV_COST_, with higher (better) Environmental Mastery, Personal Growth, Personal Relations, Purpose in Life, Self-acceptance and total score on the Psychological Wellbeing Scale associated with preservation of gait under dual-task conditions (p<0.05). Similarly, CV_COST_ was inversely associated with Duke Social Support Index Scale (DSSIS), indicating that the lower level of, and satisfaction with, social support was associated with a greater gait impairment during dual-task conditions (p<0.05).

### Factors associated with changes in fractal scaling exponent of gait during dual-tasking (DFA_*COST*_)

For simplicity, significant associations are presented in Table 2. A complete set of results can be seen in the Supplemental material. Contrary to our hypotheses, cognition and psychosocial function were not associated with DFA_COST_ under dual-task conditions (p>0.05). Notably, as anticipated, greater left (r=0.23, p=0.026) and total (r=0.25, p=0.015) posterior cingulate cortex thickness as well as better performances in the 6MWT (r=0.25, p=0.025) and aerobic capacity (r=0.24, p=0.033) were related to preservation of gait during dual-tasking. However, DFA_COST_ was unrelated to lower body strength (r=0.25, p=0.063), whole body strength (r=0.25, p=0.079), static balance time (r=0.18, p=0.075) or tandem walk score (r=-0.12, p=0.254).

## DISCUSSION

The primary finding from this novel investigation was that in older adults with MCI, resting cognitive performance on a letter fluency task was *preserved* under dual-task conditions, whereas a significant *worsening* of gait dynamics, both in the magnitude (CV) and time course (DFA), was observed. This finding in adults with MCI is consistent with the concept of “posture second” that has previously been observed in healthy older adults^27^ and adults with Parkinson’s disease^28^. In adults with MCI who have a reduced cognitive reserve compared to cognitively healthy adults, the preservation of cognitive function at the expense of a stable gait may partially explain why people with MCI are at a greater risk of falling than healthy peers^5^. Furthermore, worsening of DFA was associated with lower posterior cingulate cortex thickness and aerobic and walking capacity, while worsening of stride time variability was associated with smaller hippocampal volume, static balance and psychological well-being. In contrast, single-task (seated) cognitive function predicted neither gait variability nor gait dynamic changes during dual-tasking. Although previous studies^4^ have focused on the decrements in gait under dual-task conditions, our study has also investigated for the first time the effects of dual tasking on cognitive performance in MCI, as well as the mediating role of brain morphology and physical/psychological function on gait stability in MCI.

This is the first report to our knowledge that thicker left and total posterior cingulate cortices were associated with preservation of DFA during dual-tasking. This is in agreement with previous data showing a greater dual task cost on gait speed (slowing down) is associated with lower grey matter volume of the hippocampus and cingulate cortex (including both anterior and posterior cingulate cortex) amongst other regions^12^. More recently, greater dual task cost on gait speed was associated with lower entorhinal cortex volume in older adults with MCI^11^, a region of the brain known to have structural and functional connections with the posterior cingulate cortex^29,30^. The posterior cingulate cortex contributes to motor imagery^31^, and assists in directing the focus of attention^32,33^, and is known to be one of the areas to atrophy early in the course of Alzheimer’s disease^33,34^. Posterior cingulate thickness reduction with age, MCI or dementia could theoretically reduce walking coordination and result in a more variable gait, particularly during dual-tasking.

We also report for the first time in this cohort that lower left and total hippocampal volume were associated with greater deficits in stride time variability during dual-tasking. The hippocampus contributes to rhythmicity of locomotion^35^ and is known to atrophy in individuals with MCI faster than in healthy adults^36^. Hippocampal volume has been associated with gait variability in both cognitively healthy adults, and adults with cognitive impairment^10^, but data are lacking under dual-task conditions. Our findings suggest that hippocampal atrophy associated with MCI may contribute to a diminished ability to regulate gait dynamics under the stressful condition of dual-tasking in this cohort.

Notably, we have previously shown that high intensity strength training increases posterior cingulate thickness^17^ and that this change in posterior cingulate thickness is directly associated with the cognitive benefits of the strength training^17^. While no change in the hippocampus was observed in SMART^17^ or other studies of progressive resistance training^37^, hippocampal volume has been shown to increase following aerobic exercise in adults with MCI^37^. Future investigations are needed to determine whether gait dynamics under dual-task conditions improve following structured aerobic or resistance training programs, and if any observed improvements are mediated by structural brain changes.

Higher aerobic capacity and 6-min walking distance were associated with better preservation of DFA during dual-tasking. High intensity treadmill training has been shown to improve aerobic capacity and cognition in adults with amnestic MCI^38^, whereas most other low-moderate intensity aerobic interventions have yielded non-significant findings in this cohort^39^. Progressive resistance training can also improve aerobic capacity and cognitive function, (mediated by muscle strength gains), as we have previously shown within this cohort^16^. However, given the cross-sectional nature of our analyses, reverse causality cannot be excluded. Individuals with a less variable gait may be more likely to walk more frequently and thus have a greater aerobic capacity and functional performance compared to individuals with a more variable gait pattern. Longitudinal exercise studies are required to investigate whether improved aerobic/walking capacity will also improve gait variability during single- and dual-task conditions.

Muscle strength was not related to the preservation of either DFA or stride time variability during dual-task walking. This differs from data we previously reported showing that higher muscle strength was related to lower variability measures in community-dwelling older adults^2^, and that improvements in muscle strength following a multi-modal exercise program were associated with reduced stride-time variability in older adults with functional impairment^40^. However, consistent with the current findings, others have reported that stride time variability in older adults with higher level gait disorders was not associated with muscle strength^24^, nor was step time variability improved by a muscle strengthening intervention^41^. All of the above studies only measured gait variability under single-task conditions, and thus, the potential benefit of interventions to improve muscle strength on gait variability under dual-task conditions are unknown.

The dual-task cost of stride time variability was greater in those with lower psychological well-being (SPWB) across the domains of Environmental Mastery, Personal Growth, Personal Relations, Purpose in Life and Self-acceptance. These data are in agreement with previous evidence suggesting that worse stride time variability is associated with an increased fear of falling^24^, and fear of falling itself has been previously associated with limitations in physical, mental and social functioning^42^. Additionally, MCI has been associated with reduced psychological wellbeing^43^ and increased falls^2^, supporting a potential link between worsened gait dynamics and poor psychological wellbeing, although reverse causality or bi-directional relationships cannot be ruled out. Consistent with our findings on psychological well-being, depression is a well-known risk factor for falls and hip fractures^44^, independent from anti-depressant medications, which pose additional fall risk^45^. It is important to note however that our cohort was free from clinical depression at entry into this study by design. Given the known relationship between hippocampal atrophy and depression^46^, the potential link between depressive symptoms, brain morphology and gait dynamics is an important area for future investigation in relevant high-risk cohorts with this co-morbidity.

### Limitations

As noted above, reverse causality may explain some of the study outcomes due to the cross-sectional study design. Also, the order of the COWAT assessment and dual-task walking was not randomized. The presentation of the “F” word task at rest prior to dual-tasking may have produced a learning effect, minimizing any potential to observe cognitive deficits during dual-tasking. Future studies should randomize the sequencing of single- and dual-task cognitive performance.

## CONCLUSIONS

Older adults with MCI preserved their cognitive performance at the cost of the variability and dynamics of their gait under dual-task conditions. Novel associations were observed between worsening of the fractal scaling exponent of gait and posterior cingulate cortical thickness, while worsening of stride time variability was associated with lower hippocampal volume. Better aerobic and walking capacity, psychological wellbeing and static balance were also associated with preservation gait stability during dual-tasking. Notably, *all* of these factors have previously been shown to be modifiable with robust exercise modalities in clinical trials. Longitudinal research is required to determine the extent to which gait dynamics are also modifiable, and the optimal exercise prescriptions needed to promote optimization of gait patterns, resilience to stressors, and ultimately reduce fall risk in vulnerable cohorts.

## Data Availability

Data are not available

## Acknowledgements

This work fulfilled a portion of the degree requirements for PhD for NG and CS, and Master’s Degree for TH. Donations for participant rewards were received from Gregory and Carr Funerals. We would also like to thank the extraordinary generosity and commitment of the participants and their families who devoted their time to the SMART study.

## Author Contributions

Study concept and design: TCH, RS,YM, MAFS and JMH. Acquisition of data: NG, GCW, NJ, JM, CS, MKB, NF and YW. Analysis and interpretation of data: TCH, RS, YM, MAFS and JMH. Drafting of the manuscript: TCH, RS, YM, JMH and MAFS. Critical revision of the manuscript for important intellectual content: TCH, RS, NG, GCW, NJ, JM, WW, MKB, NF, YW, HB, NS, BTB, CS, PSS, MV, YM, MAFS and JMH. Statistical analysis: TCH, RS and YM. Obtained funding: HB, WW, NS, BTB, PSS, MV, MAFS. Administrative, technical, and material support: NJ, JM, CS. Study supervision: NG, HB, PSS, MV and MAFS.

## REFERENCES

1. Verghese J, LeValley A, Hall CB, Katz MJ, Ambrose AF, Lipton RB. Epidemiology of Gait Disorders in Community-Residing Older Adults. Journal of the American Geriatrics Society. 2006;54(2):255–261. https://doi.org/10.1111/j.1532-5415.2005.00580.x

2. Callisaya ML, Blizzard L, Schmidt MD, et al. Gait, gait variability and the risk of multiple incident falls in older people: a population-based study. Age and Ageing. 2011;40(4):481–487. https://doi.org/10.1093/ageing/afr055

3. Brach JS, Studenski SA, Perera S, VanSwearingen JM, Newman AB. Gait variability and the risk of incident mobility disability in community-dwelling older adults. The Journals of Gerontology Series A: Biological Sciences and Medical Sciences. 2007;62(9):983–988. https://doi.org/10.1093/gerona/62.9.983

4. Montero-Odasso M, Muir SW, Speechley M. Dual-task complexity affects gait in people with mild cognitive impairment: the interplay between gait variability, dual tasking, and risk of falls. Arch Phys Med Rehabil. 2012;93(2):293–299. https://doi.org/10.1016/j.apmr.2011.08.026

5. Delbaere K, Kochan NA, Close JCT, et al. Mild Cognitive Impairment as a Predictor of Falls in Community-Dwelling Older People. The American Journal of Geriatric Psychiatry. 2012;20(10):845–853. https://doi.org/10.1097/JGP.0b013e31824afbc4

6. Montero-Odasso M, Bergman H, Phillips NA, Wong CH, Sourial N, Chertkow H. Dual-tasking and gait in people with mild cognitive impairment. The effect of working memory. BMC Geriatrics. 2009;9(41). https://doi.org/10.1186/1471-2318-9-41

7. Beauchet O, Annweiler C, Dubost V, et al. Stops walking when talking: A predictor of falls in older adults? European Journal of Neurology. 2009;16(7):786–795. https://doi.org/10.1111/j.1468-1331.2009.02612.x

8. Muir-Hunter SW, Wittwer JE. Dual-task testing to predict falls in community-dwelling older adults: a systematic review. Physiotherapy. 2016;102(1):29–40. https://doi.org/10.1016/j.physio.2015.04.011

9. Montero-Odasso M, Verghese J, Beauchet O, Hausdorff JM. Gait and cognition: A complementary approach to understanding brain function and the risk of falling. Journal of the American Geriatrics Society. 2012;60(11):2127–2136. https://doi.org/10.1111/j.1532-5415.2012.04209.x

10. Tian Q, Chastan N, Bair W-N, Resnick SM, Ferrucci L, Studenski SA. The brain map of gait variability in aging, cognitive impairment and dementia—a systematic review. Neuroscience & Biobehavioral Reviews. 2017;74:149–162. https://doi.org/10.1016/j.neubiorev.2017.01.020

11. Sakurai R, Bartha R, Montero-Odasso M. Entorhinal Cortex Volume Is Associated With Dual-Task Gait Cost Among Older Adults With MCI: Results From the Gait and Brain Study. The Journals of Gerontology: Series A. 2018;74(5):698–704. https://doi.org/10.1093/gerona/gly084

12. Doi T, Blumen HM, Verghese J, et al. Gray matter volume and dual-task gait performance in mild cognitive impairment. Brain Imaging and Behavior. 2017;11(3):887–898. https://10.1007/s11682-016-9562-1

13. Clegg A, Young J, Iliffe S, Rikkert MO, Rockwood K. Frailty in elderly people. The Lancet. 2013;381(9868):752–762. https://doi.org/10.1016/S0140-6736(12)62167-9

14. Gates NJ, Valenzuela M, Sachdev PS, et al. Study of Mental Activity and Regular Training (SMART) in at risk individuals: A randomised double blind, sham controlled, longitudinal trial. BMC Geriatrics. 2011;11(1):19. https://doi.org/10.1186/1471-2318-11-19

15. Fiatarone Singh MA, Gates N, Saigal N, et al. The Study of Mental and Resistance Training (SMART) study—resistance training and/or cognitive training in mild cognitive impairment: a randomized, double-blind, double-sham controlled trial. Journal of the American Medical Directors Association. 2014;15(12):873–880. https://doi.org/10.1016/j.jamda.2014.09.010

16. Mavros Y, Gates N, Wilson GC, et al. Mediation of Cognitive Function Improvements by Strength Gains After Resistance Training in Older Adults with Mild Cognitive Impairment: Outcomes of the Study of Mental and Resistance Training. Journal of the American Geriatrics Society. 2017;65(3):550–559. https://doi.org/10.1111/jgs.14542

17. Suo C, Fiatarone Singh M, Gates N, et al. Therapeutically relevant structural and functional mechanisms triggered by physical and cognitive exercise. Molecular Psychiatry. 2016;21:1633–1642. https://doi.org/10.1038/mp.2016.19

18. Petersen RC. Mild Cognitive Impairment: Clinical Characterization and Outcome. Archives of Neurology. 1999;281(19):303–308. https://doi.org/10.1001/archneur.56.3.303

19. Kaufman AS, Lichtenberger EO. Essentials: of WAIS-III assessment. New York: J. Wiley & Sons; 1999.

20. Hausdorff JM, Edelberg HK, Mitchell SL, Goldberger AL, Wei JY. Increased gait unsteadiness in community-dwelling elderly fallers. Archives of physical medicine and rehabilitation. 1997;78(3):278–283. https://doi.org/10.1016/S0003-9993(97)90034-4

21. Hausdorff JM, Ladin Z, Wei JY. Footswitch system for measurement of the temporal parameters of gait. Journal of Biomechanics. 1995;28(3):347–351. https://doi.org/10.1016/0021-9290(94)00074-E

22. Hausdorff JM, Peng CK, Ladin Z, Wei JY, Goldberger AL. Is walking a random walk? Evidence for long-range correlations in stride interval of human gait. Journal of Applied Physiology. 1995;78(1):349–358. https://doi.org/10.1152/jappl.1995.78.1.349

23. Hausdorff JM, Purdon PL, Peng CK, Ladin Z, Wei JY, Goldberger AL. Fractal dynamics of human gait: stability of long-range correlations in stride interval fluctuations. Journal of Applied Physiology. 1996;80(5):1448–1457. https://doi.org/10.1152/jappl.1996.80.5.1448

24. Herman T, Giladi N, Gurevich T, Hausdorff JM. Gait instability and fractal dynamics of older adults with a “cautious” gait: why do certain older adults walk fearfully? Gait & Posture. 2005;21(2):178–185. https://doi.org/10.1016/j.gaitpost.2004.01.014

25. Suo C, Leon I, Brodaty H, et al. Supervisory experience at work is linked to low rate of hippocampal atrophy in late life. Neuroimage. 2012;63(3):1542–1551. https://10.1016/j.neuroimage.2012.08.015

26. Desikan RS, Segonne F, Fischl B, et al. An automated labeling system for subdividing the human cerebral cortex on MRI scans into gyral based regions of interest. Neuroimage. 2006;31(3):968–980. https://10.1016/j.neuroimage.2006.01.021

27. Corp DT, Youssef GJ, Clark RA, et al. Reduced motor cortex inhibition and a ‘cognitive- first’ prioritisation strategy for older adults during dual-tasking. Experimental Gerontology. 2018;113:95–105. https://doi.org/10.1016/j.exger.2018.09.018

28. Bloem BR, Grimbergen YAM, van Dijk JG, Munneke M. The “posture second” strategy: A review of wrong priorities in Parkinson’s disease. Journal of the Neurological Sciences. 2006;248(1-2):196-204. https://doi.org/10.1016/j.jns.2006.05.010

29. Buckner RL, Andrews-Hanna JR, Schacter DL. The brain’s default network: anatomy, function, and relevance to disease. Ann N Y Acad Sci. 2008;1124:1–38. https://10.1196/annals.1440.011

30. Greicius MD, Supekar K, Menon V, Dougherty RF. Resting-State Functional Connectivity Reflects Structural Connectivity in the Default Mode Network. Cerebral Cortex. 2008;19(1):72–78. https://doi.org/10.1093/cercor/bhn059

31. Rosano C, Aizenstein H, Brach J, Longenberger A, Studenski S, Newman AB. Special ArticleGait Measures Indicate Underlying Focal Gray Matter Atrophy in the Brain of Older Adults. The Journals of Gerontology: Series A. 2008;63(12):1380–1388. https://doi.org/10.1093/gerona/63.12.1380

32. Leech R, Kamourieh S, Beckmann CF, Sharp DJ. Fractionating the Default Mode Network: Distinct Contributions of the Ventral and Dorsal Posterior Cingulate Cortex to Cognitive Control. The Journal of Neuroscience. 2011;31(9):3217–3224. https://doi.org/10.1523/JNEUROSCI.5626-10.2011

33. Leech R, Sharp DJ. The role of the posterior cingulate cortex in cognition and disease. Brain. 2013;137(1):12–32. https://10.1093/brain/awt162

34. Buckner RL, Snyder AZ, Shannon BJ, et al. Molecular, Structural, and Functional Characterization of Alzheimer’s Disease: Evidence for a Relationship between Default Activity, Amyloid, and Memory. The Journal of Neuroscience. 2005;25(34):7709–7717. https://doi.org/10.1523/JNEUROSCI.2177-05.2005

35. Zimmerman ME, Lipton RB, Pan JW, Hetherington HP, Verghese J. MRI- and MRS-Derived Hippocampal Correlates of Quantitative Locomotor Function in Older Adults. Brain research. 2009;1291:73–81. https://doi.org/10.1016/j.brainres.2009.07.043

36. Leal SL, Yassa MA. Perturbations of neural circuitry in aging, mild cognitive impairment, and Alzheimer’s disease. Ageing Research Reviews. 2013;12(3):823–831. https://doi.org/10.1016/j.arr.2013.01.006

37. ten Brinke LF, Bolandzadeh N, Nagamatsu LS, et al. Aerobic exercise increases hippocampal volume in older women with probable mild cognitive impairment: a 6- month randomised controlled trial. British Journal of Sports Medicine. 2015;49(4):248–254. https://doi.org/10.1136/bjsports-2013-093184

38. Baker LD, Frank LL, Foster-Schubert K, et al. Effects of Aerobic Exercise on Mild Cognitive Impairment: A Controlled Trial. Archives of neurology. 2010;67(1):71–79. https://doi.org/10.1001/archneurol.2009.307

39. Gates N, Singh MAF, Sachdev PS, Valenzuela M. The effect of exercise training on cognitive function in older adults with mild cognitive impairment: a meta-analysis of randomized controlled trials. The American Journal of Geriatric Psychiatry. 2013;21(11):1086–1097. https://doi.org/10.1016/j.jagp.2013.02.018

40. Hausdorff JM, Nelson ME, Kaliton D, et al. Etiology and modification of gait instability in older adults: a randomized controlled trial of exercise. Journal of Applied Physiology. 2001;90(6):2117–2129. https://doi.org/10.1152/jappl.2001.90.6.2117

41. Schwenk M, Zieschang T, Englert S, Grewal G, Najafi B, Hauer K. Improvements in gait characteristics after intensive resistance and functional training in people with dementia: a randomised controlled trial. BMC Geriatrics. 2014;14(1):73. https://doi.org/10.1186/1471-2318-14-73

42. Meulen E, Zijlstra G, Ambergen T, Kempen GI. Effect of FallLRelated Concerns on Physical, Mental, and Social Function in CommunityLDwelling Older Adults: A Prospective Cohort Study. Journal of the American Geriatrics Society. 2014;62(12):2333–2338. https://doi.org/10.1111/jgs.13083

43. Gates N, Valenzuela M, Sachdev PS, Fiatarone Singh MA. Psychological well-being in individuals with mild cognitive impairment. Clinical Interventions in Aging. 2014;9:779–792. https://doi.org/10.2147/CIA.S58866

44. Whooley MA, Kip KE, Cauley JA, et al. Depression, Falls, and Risk of Fracture in Older Women. Archives of Internal Medicine. 1999;159(5):484–490. https://doi.org/10.1001/archinte.159.5.484

45. Kvelde T, Lord SR, Close JCT, et al. Depressive symptoms increase fall risk in older people, independent of antidepressant use, and reduced executive and physical functioning. Archives of Gerontology and Geriatrics. 2015;60(1):190–195. https://doi.org/10.1016/j.archger.2014.09.003

46. Sheline YI, Wang PW, Gado MH, Csernansky JG, Vannier MW. Hippocampal atrophy in recurrent major depression. Proceedings of the National Academy of Sciences. 1996;93(9):3908–3913. https://doi.org/10.1073/pnas.93.9.3908

